# Deep sleep homeostatic response to naturalistic sleep loss

**DOI:** 10.1101/2024.10.19.24315819

**Authors:** Balaji Goparaju, Sharon Ravindran, Matt T. Bianchi

## Abstract

**Introduction:** Investigations of sleep homeostasis often involve tightly controlled experimental sleep deprivation in service of understanding mechanistic physiology. The extent to which the deep sleep response to recent sleep loss occurs in naturalistic settings remains under-studied. We tested the hypothesis that a homeostatic increase in deep sleep occurs on the night following occasional short duration nights that arise in naturalistic settings.

**Methods:** We analyzed sleep staging data in participants who provided informed consent to participate in the Apple Heart and Movement Study and elected to contribute sleep data. The analysis group included n=44,564 participants with at least 30 nights of sleep staging data from Apple Watch, from November 2022 to May 2023, totaling over 5.3 million nights.

**Results:** Short nights of sleep that were >=2 hours shorter than each participant’s median sleep duration occurred at least once in 92.9% of the cohort, most often in isolation (<7% of instances were consecutive short nights), and with a median duration of just over 4 hours. We observed that the amount of deep sleep increased on the subsequent night in proportion to the amount of sleep loss on the preceding short night, in a dose response manner for short night definitions ranging from 30 minutes to >=3 hours below the within-participant median sleep duration. Focusing on short nights that were at least 2 hours below the median duration, we found that 58.8% of participants showed any increase in subsequent deep sleep, with a median increase of 12% (absolute increase of 5 minutes). In addition, the variability in deep sleep after short nights markedly increased in a dose response manner. The deep sleep homeostatic response showed little correlation to sleep duration, timing, consistency, or sleep stages, but was inversely correlated with deep sleep latency (Spearman R = -0.28).

**Conclusion:** The results provide evidence for homeostatic responses in a real-world setting. Although the deep sleep rebound amounts are modest, naturalistic short nights are a milder perturbation compared to experimental deprivation, and reactive behaviors potentially impacting sleep physiology are uncontrolled. The marked increase in variability of deep sleep amount after short nights may reflect unmeasured reactive behaviors such as caffeine or napping, which exert opposing pressures on deep sleep compared to the homeostat. The findings illustrate the utility of longitudinal sleep tracking to assess real-world correlates of sleep phenomenology established in controlled experimental settings.

## 1. Introduction

For over 40 years, sleep has been conceptualized as a two process model, with a circadian system controlling the timing of sleep, and a homeostatic system that regulates sleep drive in response to recent sleep history, manifesting for example as an increased sleep drive to counterbalance recent sleep loss^1^. Within the umbrella concept of sleep homeostasis, also known as Process S^2,3^, the responsiveness of deep sleep in particular to recent sleep-wake history has been extensively studied from the perspective of signal processing of slow wave electroencephalography (EEG) data, and also through the lens of scored sleep staging (N3; also known as deep sleep or slow wave sleep). The most robust experimental demonstration of the homeostatic response derives from total sleep deprivation in a laboratory setting, which results in rebound of deep sleep on the subsequent sleep opportunity. Variations of the classic laboratory deprivation protocols include other techniques to examine sleep homeostasis, such as in-home deprivation^4^, repeated restriction (partial deprivation) over multiple nights^5^, and selective disruption of deep sleep without reducing total sleep time, for example via targeted acoustic stimuli^6^. Although the homeostatic response to total deprivation is well described, individual variability in physiological response is also increasingly appreciated^7–9^. Even under controlled experimental settings of total deprivation with healthy subjects, the inter-individual variability of deep sleep response is evident in the prominent standard deviation values of the reported changes^10,11^. Variability in homeostatic response is also evident in the setting of repeated sleep restriction or selective deep sleep disruption, where some studies show deep sleep homeostatic increases^6,12^, while others show little or no deep sleep rebound^5,13–19^. These findings raise the question of how much homeostatic physiology might be evident in more realistic sleep patterns of sleep loss.

Highly controlled experimental settings serve an important purpose of describing and testing fundamental physiological processes while minimizing confounders that can obscure mechanistic inferences. However, the extent to which deep sleep exhibits a homeostatic response to a short night of sleep in natural real-world settings remains uncertain. For example, in experimental studies, a person’s environment (especially light) and behavior (e.g., exercise, naps, caffeine, or alcohol), which might impact sleep processes, are tightly controlled before, during, and after the deprivation intervention (e.g., see ^10,20^). Real-world sleep loss is expected to be quite distinct from experimental sleep restriction or deprivation, regarding not only the amount of loss, but also the variety of intrinsic and extrinsic factors that might influence the risk of having a night with sleep loss, the next-day countermeasures in reaction to sleep loss, the timing and opportunity of subsequent sleep, and other potential behavioral confounders.

The current work tests the hypothesis that deep sleep homeostasis can manifest in real-world settings as directional changes in the amount of deep sleep, as measured by a validated smart watch feature^21^, after short duration nights that naturally occur in a large real-world cohort with longitudinal sleep data.

## 2. Methods

### 2.1 AHMS overview

This study included participants who consented to participate in the Apple Heart and Movement Study (AHMS)^22,23^, and opt in to sharing HealthKit data streams including sleep tracking data. The Study was approved by the Advarra Central Institutional Review Board, and registered to ClinicalTrials.gov (ClinicalTrials.gov Identifier: NCT04198194). Informed consent is provided within the Apple Research app. A variety of data types including objective sleep and exercise data, written to HealthKit, are collected in the AHMS. Participants opt in to sharing data and can withdraw from the study at any time.

### 2.2 Study population

The population consists of adults living in the United States, and who own an iPhone and an Apple Watch. Sleep tracking is not required or incentivized in the study. The inclusion criteria for the current work consisted of those with at least 30 nights of sleep staging data from native Apple Watch (“first party”) sleep tracking. The analysis group included n=44,564 participants with at least 30 nights with first party sleep staging data from Apple Watch in the time window from Nov 1, 2022 to May 1, 2023.

### 2.3 Data Types

Sleep data was obtained from HealthKit, and the total sleep duration was computed for each night when the data source was Apple Watch first party sleep tracking (Watch OS 9, iOS 16, and later). The sleep feature includes a validated algorithm^21^ that classifies every 30 seconds of a recording session into one of four states: awake, Core Sleep, Deep Sleep, or REM sleep. Deep sleep corresponds to stage N3 in the scoring rubric of the American Academy of Sleep Medicine. Core sleep corresponds to the combination of stages N1 and N2.

### 2.4 Operationalizing short night labels at the participant level

Each participant was assessed for instances of short nights, defined in several ways. The primary analysis specified different thresholds of sleep duration that are lower than the median of all available nights for each participant. The thresholds considered for dose-response analysis were 0.5 to 1 hour, 1-2 hours, 2-3 hours, and 3+ hours lower than the median sleep duration computed within each participant. Additional analysis was undertaken using a single threshold of 2+ hours below the median duration within each participant. We performed parallel analyses by defining short nights using a relative measure of variance (z-score), allowing each participant to have a personalized threshold of what is short that depended on their night-to-night variability in sleep duration.

To be considered for the homeostatic response analysis, any night flagged as short must be followed immediately by a subsequent night that contained sleep staging data. Deep sleep after short nights was quantified in absolute (minutes) and relative (percent) terms. Short nights followed by a night without sleep data (i.e., sleep data was missing) were ignored, which amounted to 34% of the short nights using the 2 hour threshold. This degree of missingness is similar to the overall availability of sleep data per participant in this data set (73.3% of nights having sleep data available per participant, with interquartile range of 51.7% to 88.6%).

### 2.5 Analysis

Statistical analyses and plotting were done in Python^24–26^. Shuffling analyses were performed by randomizing the dates of sleep data within-participant, to remove the temporal association of short nights and subsequent nights of sleep, as a form of null-hypothesis testing. Where P-values are incorporated, we used a 0.05 threshold for significance; multiple comparison adjustments were not made.

## 3. Results

### 3.1. Cohort characteristics

**Table 1** shows the demographics and sleep characteristics of the cohort. For the distributions of sleep stages, a median value is computed for each participant, from which the population computations (median and interquartile range (IQR)) are reported for continuous variables in the table.

**Table 1.** Cohort characteristics.

| Metric | Value |
| --- | --- |
| N | 44,564 |
| Age (years) | 44 (35, 55) |
| Sex (%) | 68.1% M, 31.9% F |
| BMI (kg/m<sup>2</sup>) | 27.4 (24.1, 31.8) |
| TST (minutes) | 403 (370, 439) |
| Core (minutes) | 272 (246, 303) |
| REM (minutes) | 83 (68, 97) |
| Deep (minutes) | 45 (36, 52) |
| WASO (minutes) | 17 (10, 28) |

Values are median (and IQR in parentheses) for each continuous variable. BMI, body mass index; REM, rapid eye movement; TST, total sleep time; WASO, wake after sleep onset

### 3.2. Index short sleep events: framing the naturalistic homeostatic response hypothesis

Figure 1. frames the differences between experimental and real-world approaches to sleep loss at a high level. Controlled experimental sleep loss may be implemented as total deprivation (**Figure 1A**), or repeated partial deprivation, also known as sleep restriction (**Figure 1B**). In each case the sleep opportunity is set by the protocol. By contrast, in the real-world, the timing and duration of sleep opportunities and sleep itself are uncontrolled and thus susceptible to influence by many intrinsic and extrinsic factors. To operationalize the concept of sleep loss in a real-world setting, we used the approach shown in **Figure 1C**. First, the median total sleep time (TST) is computed within each participant using all available nights, which we consider their “baseline” TST. Then, deviations in TST to shorter durations are ascertained relative to this personalized baseline using thresholds and/or bins to describe the degree of shortness. In this sense, the short night definition is determined in a relative manner, for each participant. For example, using a 2-hour threshold to define a short night, a participant with a median TST of 7 hours would have any night of 5 hours or less in duration flagged as short, whereas a participant with a median TST of 8 hours would have any night 6 hours or less in duration flagged as short. We also used a variance-based determination of shortness (see next section). The homeostatic response is assessed using the Deep sleep staging data from the night immediately following a short night.

**Figure 1.**
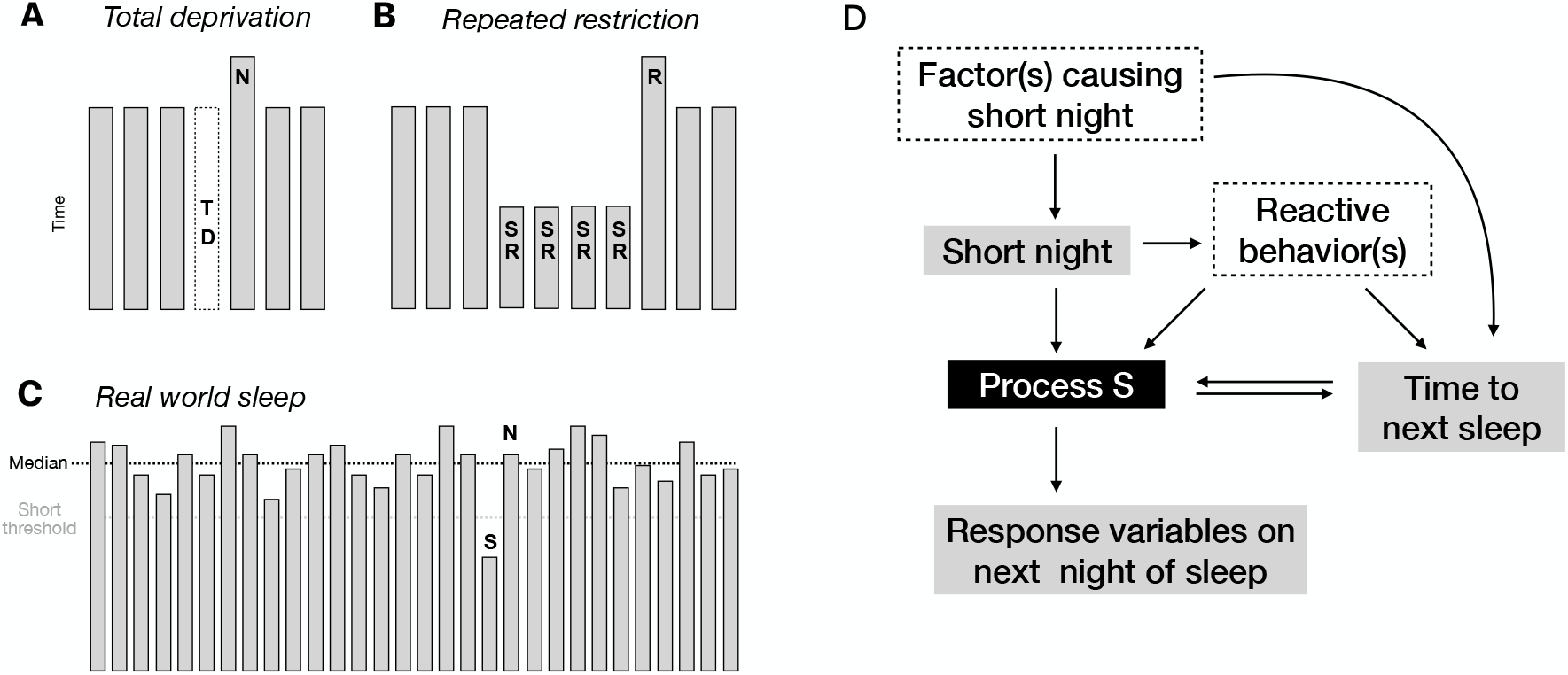
Assessing sleep homeostasis in real-world data. A. Schematic of example protocol for assessing total deprivation (TD), where “N” denotes the next (subsequent) night after TD. B. Example of repeated sleep restriction protocol (SR), with decreased sleep opportunity. C. Schematic of real-world sleep data, aligned to the start of each night for visual convenience, to visualize the approach to computing the baseline (median; dotted line) and defining short night (S) threshold (gray dotted line), and the next night (N) analyzed for homeostatic response. D. The influence diagram shows a combination of measured and unmeasured factors associated with homeostatic response to sleep loss (Process S). Gray boxes may be measurable in naturalistic data, while dotted line boxes may or may not be. Note that certain reactive behaviors, e.g., naps and caffeine (not measured in the current study), could have counteracting impact on homeostatic response variables, e.g., by reducing deep sleep on the subsequent night.

To frame the hypothesis that naturally occurring short nights are associated with subsequent homeostatic response, an influence diagram is provided to illustrate the measured versus unmeasured factors impacting sleep (**Figure 1D**). One key unknown involves what might have caused a short night in the first place. Next, how short the night was will impact the homeostatic drive, Process S, which in turn can manifest as changes in one or more homeostatic response variables on the following night. In this wearable tracking setting, we focused on three physiological response variables: Deep sleep amount (minutes), Deep sleep latency (minutes), and TST. The elapsed time between the short night and the subsequent night has a bi-directional relationship with Process S: homeostatic pressure will tend to reduce this value (increase the probability of sleep happening earlier), but if this wake time is prolonged, this can further increase homeostatic drive. Another key category, which was not measured in this study, consists of reactive behavior(s) in reaction to a short night. Such compensatory behaviors could include, for example, caffeine intake or napping, after a short night of sleep. Each could counteract the homeostatic drive caused by the short night, and could also extend the time to start the subsequent night, which then could increase homeostatic drive. Taken together, even this simplistic influence diagram captures the complexities that may arise from a basic set of factors relevant to real-world data. The framework predicts that sleep homeostasis responses in naturalistic data will be smaller than that seen in controlled experimental settings. Furthermore, it predicts that the variability in deep sleep response will expand due to behavioral countermeasures influencing deep sleep following short nights.

### 3.3. Short nights of sleep and homeostatic response variables on subsequent nights

In this cohort, we observed 302,357 short nights, using the 2-hour threshold definition, which is 5.7% of 5.33 million nights of sleep included. At the participant level, 92.9% of participants (n=41,417) had at least one short night by the 2-hour threshold definition. Those meeting criteria were generally similar to those excluded, although many of the small differences were statistically significant owing to the large sample (**Supplemental Table 1**).

The number of short nights observed per participant is skewed with a rightward tail of progressively fewer participants showing higher numbers of short nights (**Figure 2A**). We also observed that most short nights occurred in isolation, with a long tail of a small portion of consecutive short night instances, as shown in **Figure 2B**, again using the 2-hour threshold definition of a short night. For example, there is an approximately 10x drop in probability per additional consecutive night qualifying as short, such that <7% of short nights occurred in a consecutive manner (note the logarithmic axis).

**Figure 2.**
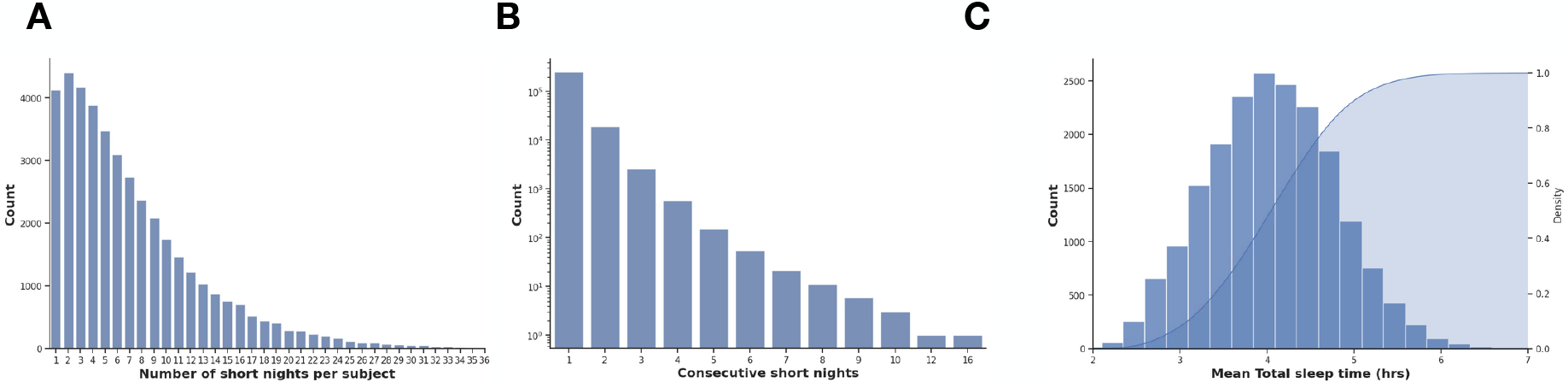
Characteristics of “short” nights using the 2-hour threshold versus the median TST. Short nights are defined here using a 2-hour threshold for comparison of each night’s TST value to within-participant median TST value. Panel A shows the distribution of number of short nights per participant, using this 2-hour threshold for labeling relative short night instances. Panel B shows the distribution of short nights in terms of isolated vs 2 or more in a row; note the logarithmic Y axis scale. Panel C is a histogram of the duration of short nights (TST), for all nights meeting the 2-hour threshold definition, with the corresponding CDF overlaid (right Y axis), showing a median of ∼4 hrs.

The distribution of TST values for short nights defined using the 2-hour threshold is shown in **Figure 2C**, with a median of 4.04 hours (IQR of 3.5 hours to 4.6 hours). The sleep staging characteristics of short nights and the following nights are given in **Supplemental Figure S1**, showing that short nights generally have less Core sleep and REM sleep (as expected given the shorter duration), and more WASO, compared to the immediately subsequent nights. At the population level, more Deep sleep was observed on subsequent nights compared to short nights. However, due to the complexities alluded to in the influence diagram of **Figure 1D**, we focused next on within-participant perspective to perform additional homeostasis response variable assessments.

At the individual level, we compared the amount of Deep sleep after a short night to the baseline Deep sleep amount, across a range of four levels of relative sleep loss: 0.5 to 1, 1-2, 2-3, and 3+ hours lower than median TST computed within each participant. We observed a small but dose-dependent increase in the amount of Deep sleep following short nights (**Figure 3**). Among short nights that were at least 2 hours below the median duration, we found that 58.8% of participants showed any increase in subsequent deep sleep, with a median increase of 12% (absolute increase of 5 minutes; IQR = 2 to 8). We also observed a similar magnitude change when homeostatic pressure is reversed: a reduction in deep sleep following particularly long nights (>=2 hours above the within-participant median; **Supplemental Figure S2**). These directional changes were small in absolute magnitude but highly statistically significant owing to the large sample size.

**Figure 3.**
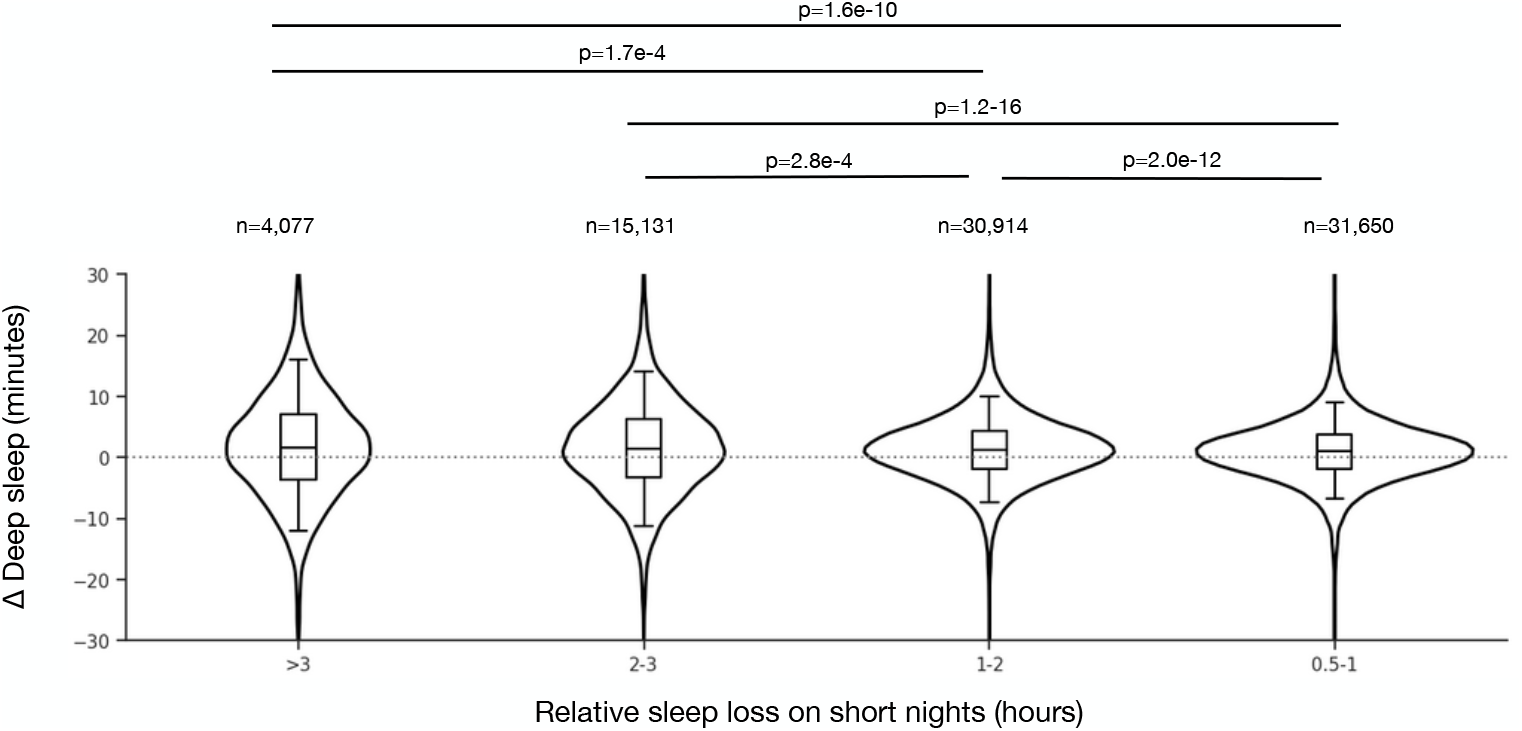
Distribution of deep sleep amounts on nights following a range of sleep loss on short nights. The X axis indicates different bins used to define relative short nights: hours below each participant’s median TST value. The Y axis shows the difference (delta, Δ, in minutes) computed within each participant as the Deep sleep on the night following a short night minus the median Deep sleep across all nights for a given participant. When more than one short night plus next-night instance was available for a given participant, the mean was used, such that one participant contributes one point in each violin group. The violin plots show the overall distributions, and each contain box plots with median, interquartile range (boxes) and 5 and 95%ile values (whiskers). The median (IQR) values from smallest to largest bins are: 1.0 (-1.8-3.7), 1.2 (-1.9-4.3), 1.4 (-3.3-6.3), 1.6 (-3.7-7.0) minutes. The sample size indicating the number of participants contributing to each threshold category is listed above the corresponding box plot (an individual can contribute to multiple violins). Mann-Whitney test results are shown for the significant comparisons, indicated by horizontal lines. The variances differed between the bins according to Levene’s test (all combinations with p< 3.9e-15).

**Figure 4.**
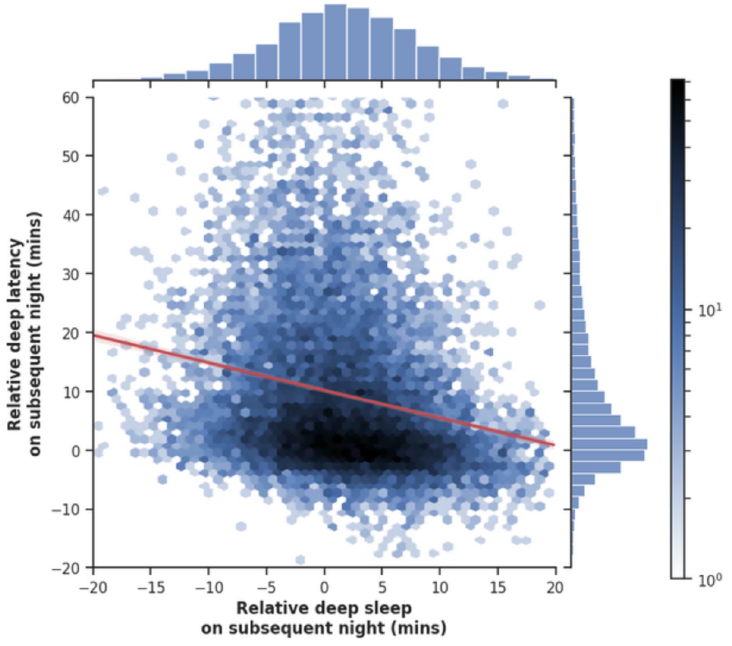
Deep sleep rebound relationship to deep sleep latency following short nights of >= 2hrs of sleep loss. The scatter plot of points is converted to a hexagonal grid with point count in each cell according to the gradient scale (log), and the corresponding marginal frequency histograms are given for each axis. When a participant has more than one short night, using the 2 hour threshold, a mean is taken. The red line is the best fit linear regression for visual convenience only. The nonparametric Spearman correlation value is -0.28 (p<0.01×10^-161^).

We also observed a clear increase in the dispersion of Deep sleep amounts across short night doses, with approximately two-fold increase in IQR with shorter nights, especially evident for the two higher sleep loss bins (**Figure 3**). For example, the IQR was 5.5 minutes for the 0.5 to 1 hour bin, compared to the IQR of 9.58 minutes for the 2-3h bin, and 10.67 minutes for the 3+ hour bin. Levene’s test for unequal variances confirmed this (less than *p*=3.9e-15 for all combinations). This pattern suggests that while on average Deep sleep amount does increase, in many instances Deep sleep amount is similar or even decreases following a short night. This variability is predicted by the potential for reactive behaviors shown **Figure 1D**.

The TST change after short nights showed a similar dose-dependent increase in dispersion (**Supplemental Figure S3**). For example, the IQR was 0.4 hours for the 0.5 to 1 hour bin, compared to the IQR of 0.85 hours for the 2-3h bin, and 1.08 hours for the 3+ hour bin.

We conducted several additional analyses to further assess the homeostatic response. First, as Deep sleep latency may be a marker of homeostatic drive, we compared this value to the Deep sleep amount, following short nights. We observed a negative correlation (R = -0.28) between the Deep sleep latency and the change in Deep sleep amount following short nights (**Supplemental Figure S4**), suggesting that nights with larger Deep sleep rebound also had shorter Deep sleep latency, that is, both moving in the direction of homeostatic prediction. Next, we repeated the dose-response analysis with two variations: quantifying the Deep sleep rebound as a percent of baseline (instead of an absolute value of minutes), and defining sleep loss relative to each participant’s nightly TST variability via bins based on z-score scaled shortness (instead of an absolute value of hours of sleep loss).

These variations showed similar dose-response relationships of magnitude and dispersion of deep sleep response (**Supplemental Figure S5 and S6**).

Given the heterogeneity of homeostatic response to sleep loss in this cohort, we sought to explore potential correlates of the response. Because the large sample sizes involved can yield significance even for very small relationships, in this exploratory analysis we instead pre-specified a Spearman R value cutoff of 0.1 as relevant, even though smaller R values can reach significance. With this threshold, we found no correlation of Deep sleep rebound with age, sex, sleep duration, sleep timing, sleep consistency (measured as the SD of duration and of start time), or sleep stage amounts (data not shown).

## 4. Discussion

This study demonstrated evidence of Deep sleep homeostasis response to episodic sleep loss using a validated consumer sleep tracking wearable in a naturalistic setting. The dose-dependent Deep sleep response and the correlation with Deep latency response support the hypothesis that some degree of physiological homeostasis occurs following naturalistic sleep loss. The increased dispersion of Deep duration and TST is consistent with a combination of individual heterogeneity in homeostatic response as well as plausible impact of unmeasured reactive behaviors predicted to counteract homeostatic drive. Although the Deep sleep rebound is small in absolute terms, the short nights observed here are relatively minor deviations from habitual sleep as compared to total deprivation experiments in which homeostatic response is most robustly observed. In fact, the naturalistic sleep loss in the current study is arguably minor even compared to sleep restriction experiments (often 4 hours less than baseline) wherein the homeostatic response is highly variable if seen at all, even in controlled settings. The naturalistic setting also does not control for the myriad factors that can influence sleep habits, including potential countermeasures (e.g., caffeine, naps) reacting to occasional short nights, which are predicted to temper the Deep sleep response and also broaden its variance – both of which are observed in the current study. Taken together, the findings illustrate the utility of longitudinal consumer sleep tracking to assess real-world correlates of sleep phenomenology established in controlled experimental settings, and set the stage for improved understanding of individual differences in physiology and physiology-behavior interactions in ecologically relevant contexts. Such translations are becoming increasingly relevant, as wearables provide methods to test the ecological validity of physiological hypotheses spanning diverse aspects of health in the real world.

### 4.1 Sleep homeostasis assessed under experimental perturbations

The classic approach of total sleep deprivation in highly controlled experimental conditions, often in small samples with strict inclusion criteria, demonstrates robust deep sleep rebound response during subsequent sleep. This homeostatic rebound is typically in the range of 30-60 minutes of excess deep sleep above baseline values^10,12,27,28^, although sometimes the deep sleep rebound after total deprivation is negligible^14^. In total deprivation studies, the standard deviation of deep sleep rebound is typically around 50% of the deep sleep rebound values, a reminder of the individual variability in homeostatic physiology, which is increasingly recognized even under controlled circumstances^8,9^.

Less robust changes in deep sleep are seen in experimental protocols of partial deprivation, also known as sleep restriction protocols, which reduce the sleep opportunity (time in bed), often by 2-4 hours below habitual sleep duration, for several nights in a row. In this repeated sleep restriction setting, the deep sleep homeostatic response is more variable and of smaller magnitude than that seen in total deprivation experiments. For example, small increases in deep sleep (approximately 13 minutes) were seen in a repeated sleep restriction of 8-hour sleepers to roughly five hours of sleep per night^18^, while somewhat larger deep sleep responses were observed in another study of four-hour restrictions^12^. Other studies did not show a deep sleep homeostatic response to partial restriction ranging from four to six hours of sleep opportunity, including work by Skorucaket al^13^, Van Dongen et al^5^, Belenky et al^16^, and a field study by Horne et al^17^. Selective deep sleep deprivation via experimental interruptions, such as from acoustic stimuli, showed approximately 20 minutes of rebound deep sleep in Ferrara et al^6^ and in Dijk et al^15^. In two other studies, deep sleep was unchanged early in the repeated restriction protocol, but showed increased amounts on the third night of restriction^19^, or after 4 nights^29^. Interestingly, Klerman et al. showed that restricting sleep duration, without extended time awake, did not alter slow wave sleep amounts^30^, suggesting that time awake may play an important role independent of sleep duration changes. In the current cohort, we observed a small but significant increase in deep sleep after two consecutive nights (data not shown). Deep sleep latency in particular has not been as well studied as a marker of homeostasis, in comparison to the efforts studying the amount of deep sleep (e.g., see^10,31,32^).

### 4.2 Real-world patterns of sleep loss

Unlike the controlled experimental settings described above, which are critical for mechanistic discovery, real-world sleep occurs within a larger context of behaviors, with direct and indirect influences on sleep, including behavioral reactions and countermeasures following recent sleep loss (**Figure 1D**). Many behavioral and lifestyle factors have been studied in the context of deep sleep in particular, including naps^33^, caffeine^34,35^, alcohol^36^, nicotine^37,38^, diet^39^, exercise^40^, as well as medications^41,42^. Certain countermeasures such as caffeine or napping, or going to bed earlier after a short night, might actually reduce homeostatic drive and thus deep sleep amount, on the subsequent night. Individual variability in the presence and chronicity of these factors, as well as individual differences in the vulnerability of sleep stages to these factors, adds uncertainty when assessing sleep in ecologically relevant settings. The impact of a counter-measure could even plausibly exceed the magnitude of the normal physiological homeostatic drive on deep sleep, creating a paradoxical reduction in deep sleep after a short night in some instances. This range of pressures on deep sleep amount following short nights predicts broader variance in deep sleep response, which was clearly evident in the current study. Uncertainty from such heterogeneity might be mitigated through protocols controlling behavior (which then reduces ecological validity), by extensive capture of potential confounders (challenging for longitudinal designs), or by large sample sizes (as in this study).

In addition to behavioral and external factors influencing sleep architecture, demographics can impact deep sleep amounts, contributing to individual variability in deep sleep and potentially its homeostatic dynamics as well^10,43,44^. Clinical conditions could also contribute variability. For example, chronic insomnia might be associated with impaired deep sleep homeostatic response^45^ (but see ^46^). Sleep apnea could impact sleep staging, and treatment of sleep apnea with positive airway pressure might result in rebound increases in deep sleep (as well as REM sleep)^47^, though the extent to which this occurs in naturalistic settings of nightly variability in adherence remains to be studied. Disentangling individual differences from inherent versus behavioral factors, and thereby improve our understanding of sleep health and its relation to overall health. Further recognizing the advantages and limitations across the spectrum of sleep assessment methods is crucial for advancing a wide spectrum of approaches to understanding sleep health^48^.

The observation that most short nights occurred in isolation might suggest at least some capacity for catch-up sleep. In that regard, although TST itself does not exhibit rebound after short nights in this cohort, TST did exhibit a similar doubling of variance, like deep sleep, after short nights. The expanded variance is in line with the speculation that concomitant unmeasured factors influence the physiological consequences of short nights. The lack of a net directional effect of short nights on TST implies that such factors on average balance out homeostatic pressure to increase TST, whereas the net impact on deep sleep is positive (i.e., net rebound) at the population level. Isolated short nights could also imply that, in this cohort at least, factors leading to sleep loss are themselves more likely to be transient. The time elapsed since awakening from the short night and starting the subsequent night was also broadly distributed (R^2^ of <0.01; data not shown), suggesting this variable was not a dominant determinant of deep sleep rebound.

### 4.3 Limitations

The current study involved a large and longitudinal sleep dataset, yet the extent to which the results generalize to other populations, or to other sleep tracking technologies, remains uncertain. Although the Watch sleep staging algorithm has been validated^21^, it is a peripheral estimate of sleep and thus the study design does not include traditional EEG markers of sleep homeostasis, such as slow wave dynamics. The effects of plausible compensatory behaviors such as naps and caffeine were not assessed, and thus remain speculative. Despite these limitations, the observation of directional changes in deep sleep after relative sleep loss is an encouraging ecological validation of using Apple Watch staging algorithm to test predictions inspired by foundational physiological principles.

## Data Availability

The aggregated data that support the findings of this study can be made available on request from the corresponding author (M.B.). Request for data will be evaluated and responded to in a manner consistent with the specific language in the study protocol and informed consent form.

## Acknowledgements

The authors acknowledge the important data contributions provided by all participants in the Apple Heart & Movement Study, without whom this research would not be possible. We also thank the following individuals for their thoughtful discussion, helpful guidance, and other valuable efforts in support of this work: Simon Saffer, Tom Furman, Callum McRae, Angela Spillane, Glen DePalma, Ian Shapiro, Asha Chesnutt, Jonathan Varbel, Jen Block, Laura Rhodes. The Apple Heart & Movement Study receives funding from Apple, Inc. and the American Heart Association.

## Supplemental Materials

**Figure S1.**
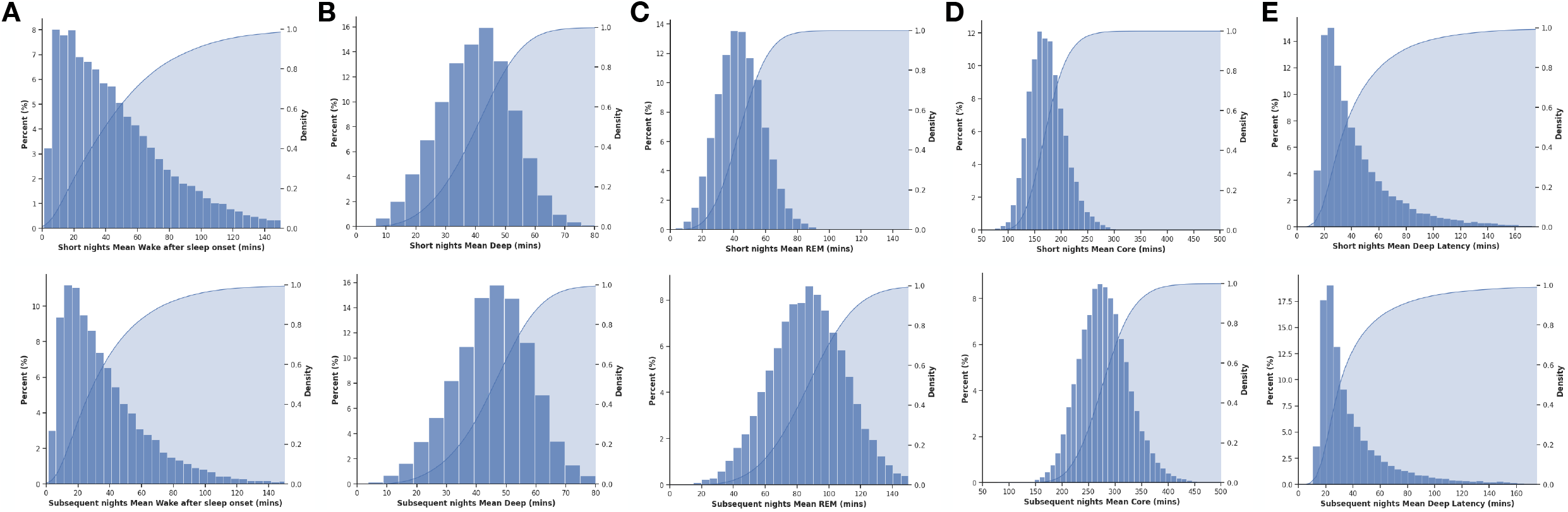
Distribution of sleep stage amounts on short nights and next nights. The top row refers to short nights, using the 2-hour threshold within-participant versus their median TST. The bottom row refers to the subsequent nights, after the short nights. In each panel, the graph shows the distribution of a sleep metric (X axis) as a frequency histogram (left Y axis) and overlaid CDF (right Y axis), for WASO (A), Deep sleep (B), REM sleep (C), Core sleep (D), and Deep Sleep Latency (E). Participants with at least one pair of short night plus subsequent night of sleep staging data are included.

**Figure S2.**
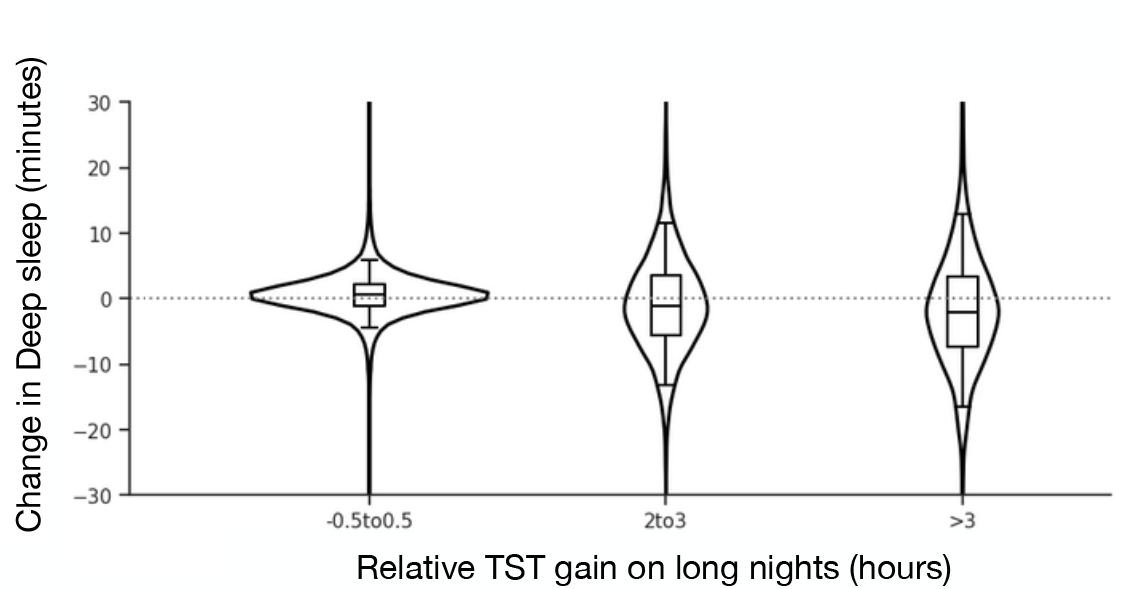
Distribution of Deep sleep changes following long nights. Violin and box plots of the Deep sleep changes observed after long nights. The bins refer to long nights defined relative to within-participant median TST. The median (IQR) values from smallest to largest bins are: 0.6 (-1.0-2.2), 1.4 (-3.3-6.3), 1.7 (-3.7-7.0) minutes. All pair-wise combinations are significantly different at p<0.015 or less (Mann Whitney). The variances differed significantly in these groups according to Levene’s test (all combinations p<0.05 or less).

**Figure S3.**
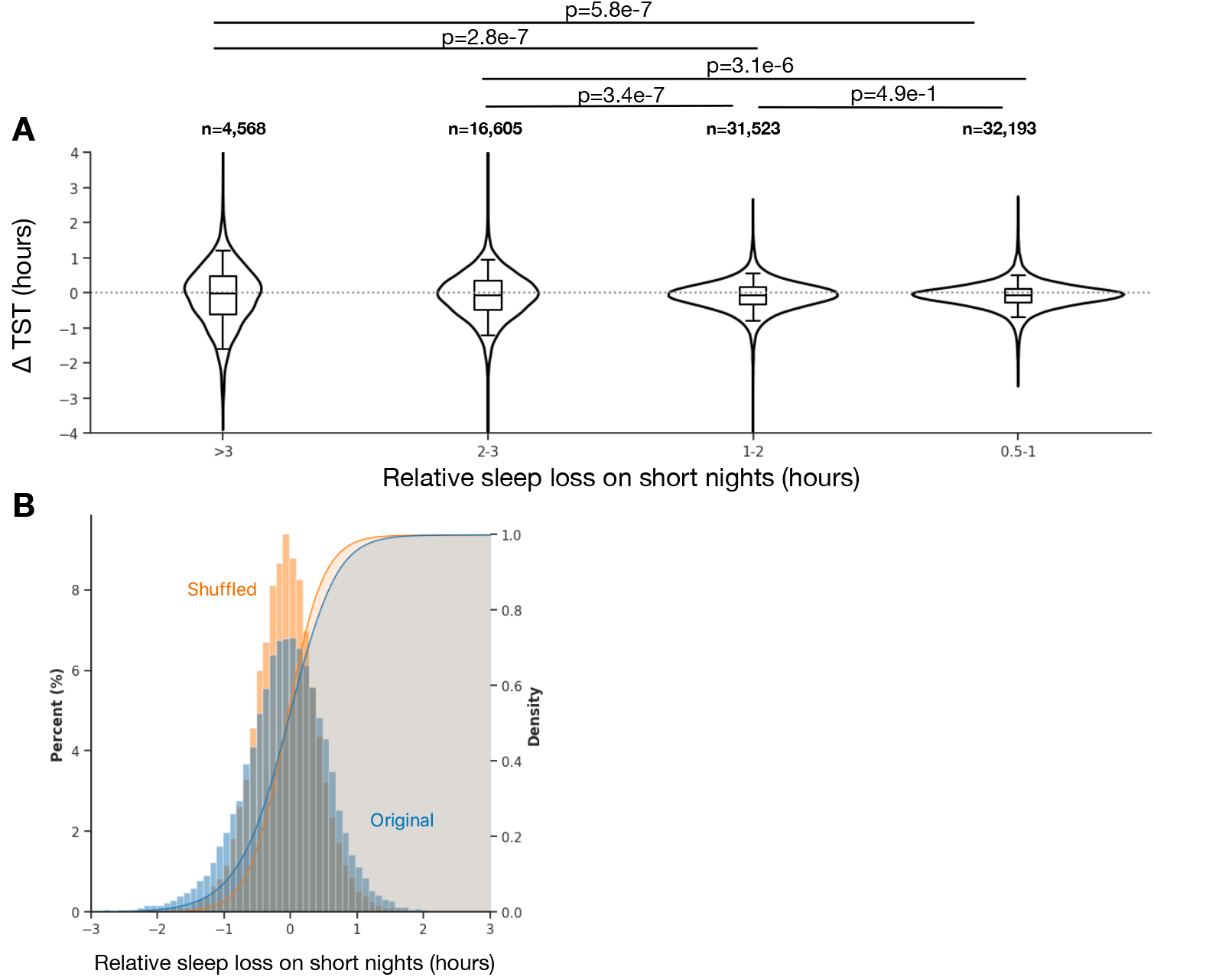
Distribution of TST changes after short nights. A. Violin and box plots of the TST changes observed following short nights across different levels of relative sleep loss. The median (IQR) values from smallest to largest bins are: -0.07 (0.28-0.12), -0.08 (-0.33-0.16), -0.05 (-0.49-0.36), -0.02 (-0.59-0.49) hours. The variances significantly differed in these groups according to Levene’s test (all combinations are p<1.9e-72). B. Histogram and overlaid CDF for the distribution of TST changes following short nights nights of >=2 hours sleep loess (blue), compared to a null distribution from date-shuffling (orange).

**Figure S5.**
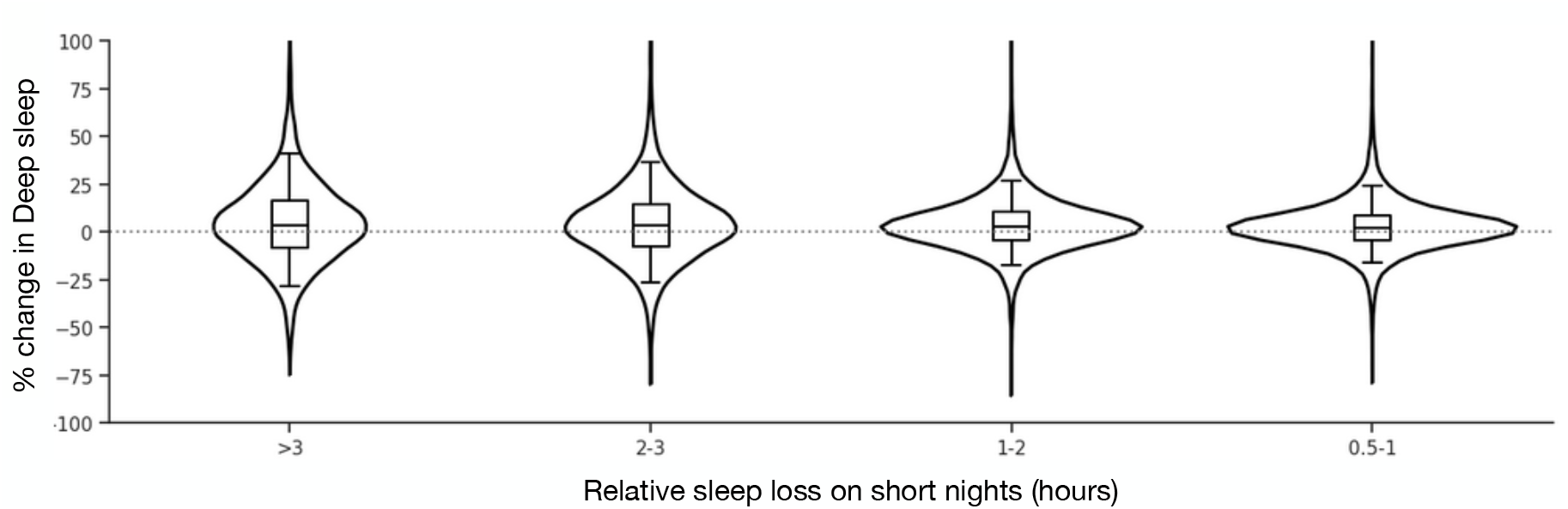
Distribution of % change in Deep sleep following short nights. Violin and box plots of the Deep sleep changes observed after different levels of relative sleep loss. The median (IQR) values from smallest to largest bins are: 2.2 (-4.2-8.9), 2.8 (-4.4-10.4), 3.3 (-7.6-14.7), 3.7 (-8.5-16.5). All pair combinations are significantly different at p<0.016 or less (Mann Whitney). The variances significantly differed in these groups according to Levene’s test (all combinations are p<7e-10).

**Figure S6.**
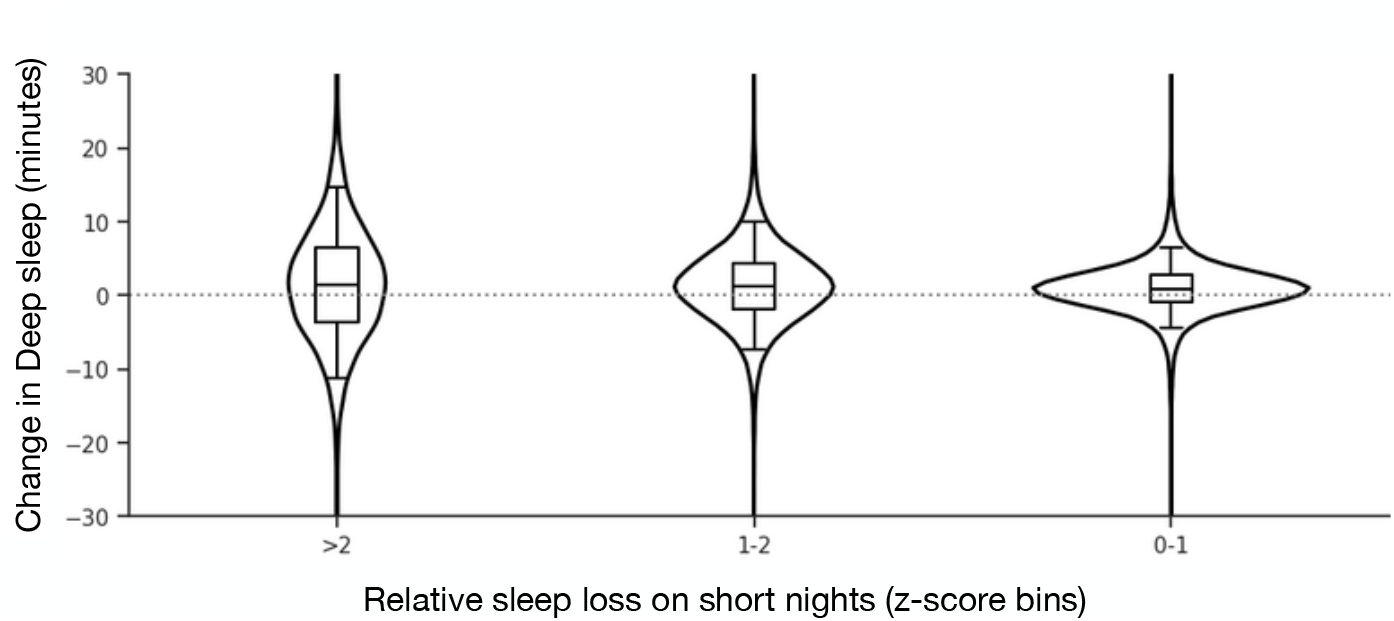
Distribution of Deep sleep changes following short nights defined by within-participant TST variance. Violin and box plots of the Deep sleep changes observed after short nights with different levels of relative sleep loss. The bins refer to short nights defined by z-score multiples based on within-participant standard deviation of nightly TST over time. The median (IQR) values from smallest to largest bins are: 0.9 (-0.9-2.7), 1.3 (-1.9-4.4), 1.5 (-3.6-6.5) minutes. All pair combinations are significantly different at p=0.001 or less (Mann Whitney). The variances differed significantly in these groups according to Levene’s test (all combinations are too small to compute.

**Supplementary Table 1.** Characteristics of participants with versus without short nights defined by a 2-hour threshold below median TST. Continuous variables are given as median (with 25th and 75th percentiles). P-values are given when lower than 0.001 (otherwise listed as NS, not-significant). BMI, body mass index. Bpm, beats per minute. IQR, interquartile range.

|  | Total | Excluded | Included | Delta<br>(Included - Excluded) |
| --- | --- | --- | --- | --- |
| N | 44564 | 3147 | 41417 | N/A |
| Age | 44 (36, 55) | 47 (38, 58) | 44 (35, 55) | -3 (1.2e-20) |
| Sex (% male) | 68.4% | 72.0% | 68.1% | -3.9 (3.8e-6) |
| BMI (kg/m²) | 27.3 (24.1, 31.7) | 26.6 (23.8, 30.8) | 27.4 (24.1, 31.8) | 0.8 (1.9e-7) |
| Active Energy (kCal) | 518, (399, 668) | 549, (422, 709) | 516 (398, 665) | -33 (3.8e-15) |
| Exercise (minutes) | 22.0, (12.0, 43.0) | 28.5, (14.0, 52.0) | 21.0 (12.0, 42.0) | -7.5 (9.4e-26) |
| rHR (bpm) | 63, (58, 69) | 62, (57, 67) | 63 (58, 69) | 1 (3.0e-18) |
| Total Sleep Time (minutes) | 403 (370, 439) | 410 (373, 443) | 403 (370, 439) | -7 (8.2e-8) |
| Core Sleep Time (minutes) | 272 (246, 303) | 274 (245, 304) | 272 (246, 303) | -2 (NS) |
| Deep Sleep Time (minutes) | 45 (36, 52) | 45 (37, 52) | 45 (36, 52) | 0 (NS) |
| REM Sleep Time (minutes) | 83 (69, 97) | 86 (72, 100) | 83 (68, 97) | -3 (2.6e-19) |
| WASO (minutes) | 17 (10, 27) | 13 (8, 21) | 17 (10, 28) | 4 (1.1e-74) |

